# Polygenic risk scores for incident dementia in the Multi-Ethnic Study of Atherosclerosis

**DOI:** 10.1101/2025.03.05.25323412

**Authors:** Diane Xue, Elizabeth E. Blue, Tamar Sofer, Timothy M. Hughes, Jerome I. Rotter, Alison E. Fohner

## Abstract

Over 75 Alzheimer’s disease (AD) and dementia-associated variants have been identified through genome-wide association studies, but the utility of polygenic risk scores (PRS) for predicting AD and dementia in diverse and admixed populations remains unclear. We compared how PRS approaches differing in *p*-value thresholds, variant weights, and source ancestry perform in predicting dementia in 6,338 African American, Chinese, Hispanic, and White individuals from the Multi-Ethnic Study of Atherosclerosis. We tested clumping and thresholding (C+T) methods with varying parameters against Bayesian approaches (PRS-CS, PRS-CSx). We compared the ability of each method to predict incident dementia in all participants and in groups stratified by self-reported race/ethnicity. We additionally analyzed performance across groups stratified by estimated proportion of non-Finnish European (NFE)-like ancestry. Including more variants does not improve performance. The PRS based on C+T method with only 15 SNPs is more strongly associated with dementia (HR_5e-08_ = 1.21, 95% CI: 1.11-1.31) than PRS derived from Bayesian models that include >800,000 SNPs (HR_CSx_ = 1.13, 95% CI: 1.04-1.23), even in populations genetically dissimilar from the source data (HR_lowNFE___5e-08_ = 1.26, 95% CI: 1.08-1.47; HR_lowNFE___CSx_ = 1.13, 95% CI: 0.96-1.32). More selective PRS models using fewer SNPs may offer better AD prediction across diverse populations.

## 1. Introduction

Dementia is a growing global health challenge, projected to affect over 150 million people worldwide by 2050(Nichols et al., 2022). Populations are aging around the world, and with nearly one third of adults over 65 dying with Alzheimer’s disease (AD) or other dementias, there is an urgent need for effective predictive tools that can aid in risk stratification and lead to more precise treatment and prevention(“2024 Alzheimer’s Disease Facts and Figures,” 2024).

AD is the most common cause of dementia and is strongly influenced by genetic variation. Less than one percent of AD cases have early-onset autosomal dominant forms of disease caused by rare coding changes in *APP, PSEN1,* or *PSEN2*(Campion et al., 1999). The vast majority of cases have a far more complex etiology. The apolipoprotein E *(APOE) ɛ4* allele is the strongest genetic risk factor for late-onset AD, but less than half of AD patients carry an *ɛ4* allele and effects are attenuated for alleles on African haplotypic backgrounds(Bertram et al., 2008; Blue et al., 2019; Pericak-Vance et al., 1991; Rajabli et al., 2018). Aside from *APOE*, dozens of loci have been found to be significantly associated with AD through genome-wide association studies (GWAS, (Andrews et al., 2023)). While these loci typically have small effect sizes, they are more common in the population, and their joint effects can place individuals at elevated genetic risk for disease.

Polygenic risk scores (PRS) based on the effects of common genetic variants have been shown to predict AD (de Rojas et al., 2021; Lambert et al., 2019; Leonenko et al., 2021). However, there is no clear consensus on the best approach for constructing PRS for AD or dementia, particularly in diverse and admixed populations. The most traditional approach for constructing PRS is clumping and thresholding (C+T), which involves initially considering all SNPs tested in a GWAS and then filtering them based on *p*-value threshold and linkage disequilibrium (LD)(Choi et al., 2020). Some previous studies have found that less stringent *p*-value thresholds, which allow for the inclusion of a larger number of SNPs in the PRS, lead to better predictive performance of AD. Escott-Price *et al*. (2015) reported that the PRS with *p*-value threshold *<* 0.5 was most strongly associated with AD(Escott-Price et al., 2015). Another study suggests that a threshold of *p <* 0.10 had optimal performance (Leonenko et al., 2019). In contrast, Zhang and colleagues found that restricting PRS to SNPs that are significantly (*p <* 1e-08) or suggestively (*p <* 3e-04) associated with AD in GWAS had better performance, implying PRS constructed using fewer than 100 SNPs can achieve superior prediction(Zhang et al., 2020). Another study found that the optimal *p*-value differed depending on the target population, ranging from *p <* 0.1 to *p <* 5e-08(Bellou et al., 2025). Notably, the comparisons discussed thus far are limited to populations with European ancestry.

Previous studies of other complex traits have shown that PRS performance deteriorates as the genetic distance between the target and GWAS training populations increases(Ding et al., 2023; Martin et al., 2019; Privé et al., 2022). Most of the risk loci discovered to be significantly associated with AD have been found in large GWAS studies of self-reported non-Hispanic White individuals who cluster with 1000 Genome (1KG) European references(Bellenguez et al., 2022; Kunkle et al., 2019). Few signals have been replicated in populations with different genetic ancestral backgrounds, including *APOE, ABCA7, TREM2, SORL1,* and *CLU*(Reitz et al., 2023). GWAS of non-European ancestry populations remain underpowered to discover risk loci with low to moderate effects, which comprise most of the AD risk loci identified thus far in large European ancestry GWAS(Xue et al., 2024). Previous studies have examined the performance of PRS for AD across populations but have limited their comparisons to C+T methods(Jung et al., 2022; Marden et al., 2014; Osterman et al., 2024; Sariya et al., 2021). We hypothesize that PRS performance across populations can be improved by using methods that retain SNPs with potential population-specific effects that would typically be excluded by pre-selected *p*-value and LD thresholds.

In this study, we assess the performance of various PRS methodologies in predicting late-onset dementia in a diverse cohort. The Multi-Ethnic Study of Atherosclerosis (MESA), a longitudinal cohort study, includes participants who self-identify as Black/African American, Chinese, Hispanic, or White. Based on GWAS of clinically ascertained AD, we compared the performance of traditional C+T methods at a range of *p*-value thresholds against Bayesian approaches (PRS-CS, PRS-CSx) using summary statistics from GWAS with differing ancestral backgrounds.

## 2. Methods

### 2.1 Study Population

MESA has been previously described in detail(Bild et al., 2002). Briefly, MESA is a prospective cohort study originally designed to study cardiovascular disease. Between 2000 and 2002, MESA recruited 6,814 Black/African American, Chinese, Hispanic/Latino, and White participants aged 45-84 from six sites in the United States: Baltimore, Maryland; Chicago, Illinois; Forsyth County, North Carolina; Los Angeles County, California; Northern Manhattan and the Bronx, New York; and Saint Paul, Minnesota. All participants were free from clinical cardiovascular disease and dementia at baseline. All participants provided written informed consent at baseline and all subsequent exams. Institutional Review Board approval was received from each of the six sites. MESA participants with imputed genotypes and information on dementia status were included in this study.

### 2.2 Inferring Global Ancestry Proportions

We estimated global ancestry proportions for all genotyped MESA participants. Global ancestry proportions are based on local ancestry estimates from RFMix2 using reference population data from the 1KG and Human Genome Diversity Project available through gnomADv3.1 as references(Auton et al., 2015; Karczewski et al., 2020; Maples et al., 2013) Samples were randomly selected from the following superpopulation groups to construct balanced sample maps: American (AMR), African (AFR), East Asian (EAS), and Non-Finnish European (NFE(Auton et al., 2015)). The genetic map with coordinates from the human reference genome GRCh38 was downloaded from the Eagle v2.4.1 package (http://data.broadinstitute.org/alkesgroup/Eagle/downloads/). Using the global ancestry proportions, participants were assigned to groups defined by low (*<* 33%), intermediate (33% - 67%), and high (>67%) NFE-like ancestry.

### 2.3 Dementia Outcome

Participants were followed up via telephone interview every 9 to 12 months for updates on hospital admissions or deaths. Dementia was identified based on a set of ICD codes at either hospitalization or death. The candidate dementia cases were identified using the following diagnosis codes: ICD-9: 290, 294, 331.0, 331.1, 331.2, 331.82, 331.83, 331.9, 438.0, and 780.93; ICD-10: F00, F01, F03, F04, G30, G31 (excluding G31.2), I69.91, and R41. The ICD code-based identification was validated against medical record text that indicates significant decline in cognitive function compared with a previous level(Fujiyoshi et al., 2017).

### 2.4 Genotyping

SNPs for all MESA participants were genotyped using the Affymetrix 6.0 SNP array. SNPs were imputed using IMPUTE version 2.2.2 and 1KG cosmopolitan phase 3 version 5 reference haplotypes. Relatedness was inferred using KING, and an unrelated subset of individuals was selected by choosing one individual at random from each first-degree related group(Manichaikul et al., 2010).

### 2.5 Calculating Polygenic Risk Scores

We compared six PRS models that excluded the *APOE* region (GRCh38 chr19: 44408822-45408822). The C+T methods were used to estimate PRS using the following *p*-value thresholds: 0.01, 1e-05, and 5e-08. SNPs were filtered based on LD *<* r^2^ = 0.01. C+T PRS were calculated using PLINK v1.90(Chang et al., 2015). After filtering based on *p*-value and LD, PRS were calculated based on the dosage of the SNP effect allele multiplied by the effect sizes. The SNPs and effect sizes for the C+T models were derived from the 2019 International Genomics of Alzheimer’s Project (IGAP) genetic meta-analysis of clinically diagnosed late-onset Alzheimer’s disease among those of European descent, which includes 21,982 cases and 41,944 controls across 46 studies(Kunkle et al., 2019). Summary statistics were obtained from the National Institute on Aging Genetics of Alzheimer’s Disease Data Storage Site (NIAGADS).

In addition to the C+T models with varying *p*-value stringency, two Bayesian models were compared: PRS-CS and PRS-CSx(Ge et al., 2019; Ruan et al., 2022). Both PRS-CS and PRS-CSx use a continuous shrinkage model that accounts for LD by tuning or shrinking the effect sizes. We did not use a separate validation set to tune parameters, instead using the -auto option for both PRS-CS and PRS-CSx. Two GWAS summary statistics were used for the PRS-CS models: the IGAP study and a cross-population GWAS of 15,579 cases and 17,690 controls that included self-reported Whites, African Americans, Japanese, and Israeli-Arabs (NG00056 (Jun et al., 2017)). PRS-CSx allows for multiple summary statistics with differing ancestral backgrounds to be used concurrently. In addition to the European ancestry IGAP study, we also included summary statistics from the African Genome Resources Panel GWAS of 2,748 cases and 5,222 controls in the same model (NG00100 (Kunkle et al., 2020)).

To calibrate the PRS for population structure, we used a previously described procedure to calculate residualized scores based on the principal components(Hao et al., 2022). We fit the raw PRS as a function of the first three principal components in non-affected individuals. We used the linear model to calculate a predicted PRS for all individuals. We then computed the residualized, population-structure adjusted PRS by calculating the difference between the raw and predicted PRS. The residualized score was then standardized based on the mean and standard deviation.

### 2.6 Statistical Analysis

Cox proportional hazards models were used to examine the association between the PRS scores and incident dementia. We additionally conducted analyses in groups stratified by self-reported race/ethnicity to examine whether the PRS models perform unequally across groups, which may exacerbate disparities. We also performed analyses stratified by quantiles of NFE-like ancestry to evaluate whether PRS performance declines as genetic distance from the training GWAS of primarily European ancestry increases. Univariate models were computed separately for each PRS method.

Harrell’s concordance (C-Index) was used to compare predictive performance. Comparisons were conducted in all participants and groups stratified by self-reported race/ethnicity. Additional comparisons were made across groups in different tertiles of European ancestry to examine if model performance was biased for those with greater proportion of European ancestry.

We computed the added predictive value of each PRS method compared to baseline models that included sex, age, and *APOE ɛ4* carrier status and used likelihood ratio tests to test the significance of improved model fit when including the PRS.

### 2.7 Sensitivity Analysis

Time to hospitalization or death due to dementia may not accurately capture dementia symptom onset or diagnosis. In addition to the comparisons of association and predictive performance based on hazard models, we also fit univariate logistic regression models to examine the association with dementia case-control status of each PRS method and calculated the area under the curve (AUC) to assess the predictive performance.

## 3. Results

### 3.1 Study population

We calculated PRS and inferred global ancestry proportions for all participants in the MESA cohort who consented to genetic analyses as part of the SNP Health Association Resource (SHARe). Of these individuals, 6,338 participants had dementia follow-up data and were included in this study. At enrollment, the mean age of the participants was 62 years. After a median follow up of 16.8 years, 560 (8.8%) incident all-cause dementia events were observed.

Complete demographic characteristics are provided in **Table 1**. The African Americans and Hispanic/Latino groups have high amounts of admixture of NFE-like and AFR-like ancestry and NFE-like, AMR-like, and AFR-like ancestry, respectively. **(Supplementary Figure 1).** The high NFE-like subgroup included 2,651 participants, intermediate NFE-like included 1,256, and low NFE-like included 2,431.

**Table 1.**
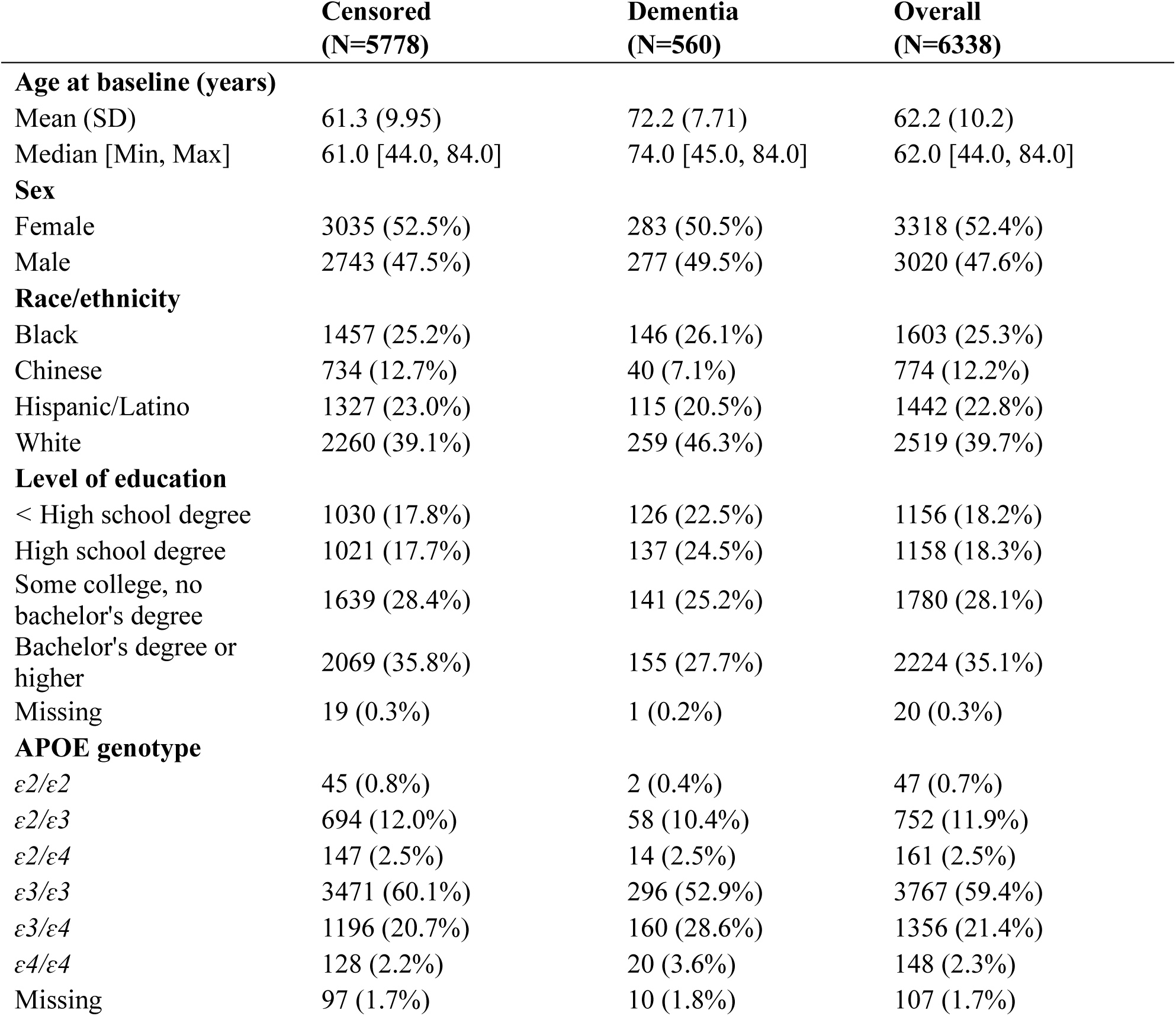
Demographic Characteristics of MESA participants at Baseline.

|  | **Censored**<br>**(N=5778)** | **Dementia**<br>**(N=560)** | **Overall**<br>**(N=6338)** |
| --- | --- | --- | --- |
| **Age at baseline (years)** |  |  |  |
| Mean (SD) | 61.3 (9.95) | 72.2 (7.71) | 62.2 (10.2) |
| Median [Min, Max] | 61.0 [44.0, 84.0] | 74.0 [45.0, 84.0] | 62.0 [44.0, 84.0] |
| **Sex** |  |  |  |
| Female | 3035 (52.5%) | 283 (50.5%) | 3318 (52.4%) |
| Male | 2743 (47.5%) | 277 (49.5%) | 3020 (47.6%) |
| **Race/ethnicity** |  |  |  |
| Black | 1457 (25.2%) | 146 (26.1%) | 1603 (25.3%) |
| Chinese | 734 (12.7%) | 40 (7.1%) | 774 (12.2%) |
| Hispanic/Latino | 1327 (23.0%) | 115 (20.5%) | 1442 (22.8%) |
| White | 2260 (39.1%) | 259 (46.3%) | 2519 (39.7%) |
| **Level of education** |  |  |  |
| < High school degree | 1030 (17.8%) | 126 (22.5%) | 1156 (18.2%) |
| High school degree | 1021 (17.7%) | 137 (24.5%) | 1158 (18.3%) |
| Some college, no<br>bachelor's degree | 1639 (28.4%) | 141 (25.2%) | 1780 (28.1%) |
| Bachelor's degree or<br>higher | 2069 (35.8%) | 155 (27.7%) | 2224 (35.1%) |
| Missing | 19 (0.3%) | 1 (0.2%) | 20 (0.3%) |
| **APOE genotype** |  |  |  |
| $\epsilon 2/\epsilon 2$ | 45 (0.8%) | 2 (0.4%) | 47 (0.7%) |
| $\epsilon 2/\epsilon 3$ | 694 (12.0%) | 58 (10.4%) | 752 (11.9%) |
| $\epsilon 2/\epsilon 4$ | 147 (2.5%) | 14 (2.5%) | 161 (2.5%) |
| $\epsilon 3/\epsilon 3$ | 3471 (60.1%) | 296 (52.9%) | 3767 (59.4%) |
| $\epsilon 3/\epsilon 4$ | 1196 (20.7%) | 160 (28.6%) | 1356 (21.4%) |
| $\epsilon 4/\epsilon 4$ | 128 (2.2%) | 20 (3.6%) | 148 (2.3%) |
| Missing | 97 (1.7%) | 10 (1.8%) | 107 (1.7%) |

### 3.2 PRS Distributions

**Table 2** outlines the number of SNPs included in each PRS model. The PRS-CSx model incorporated the largest number of SNPs (968,595) that intersect across the 1KG LD reference maps, IGAP European ancestry GWAS summary statistics, African Genome Resources Panel GWAS summary statistics, and MESA genotyped + imputation data. The PRS-CS models with the European IGAP GWAS and cross-population GWAS included 862,647 SNPs and 851,128 SNPS, respectively. In contrast, the C+T model with a stringent genome-wide significant *p*-value cutoff (*p <* 5e-08 C+T) included only 15 SNPs after filtering for LD.

**Table 2.**
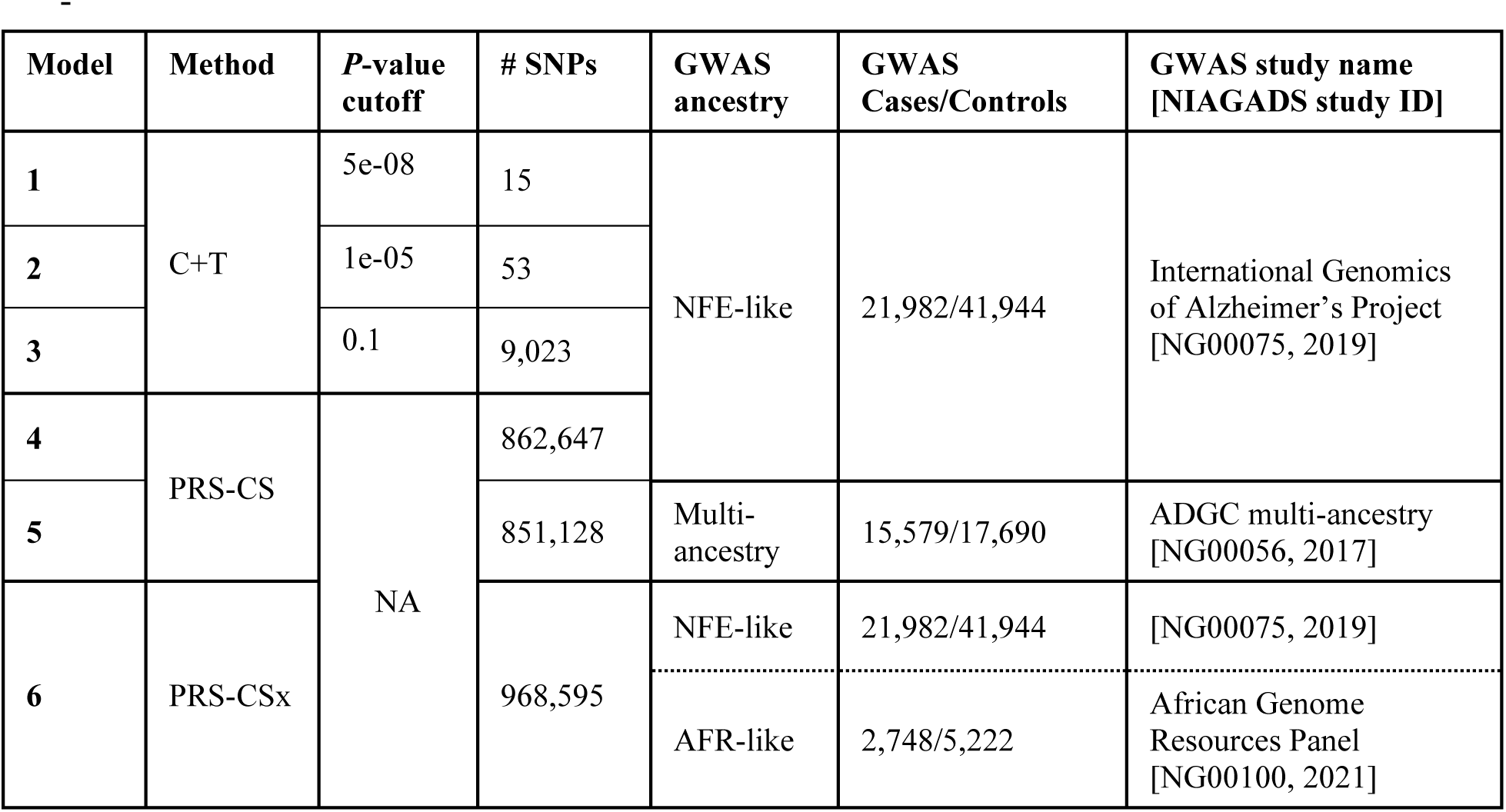
Description of PRS models. This table shows the six PRS models being compared. The models differ in computational method, *p*-value threshold and corresponding and the number of SNPs included, and/or GWAS training data. The NIAGADS study number is provided for the GWAS summary statistics used. LD reference panels were obtained from 1000 Genomes Project phase 3 samples (EUR and AFR) and used for the PRS-CS and PRS-CSx models.

For all models, marked differences in PRS distributions were observed across self-reported racial groups using raw scores, although the differences are less pronounced in the conservative C+T models. The variation was attenuated after calibrating for population structure. (**Figure 1**, **Supplementary Figure 2**).

**Figure 1.**
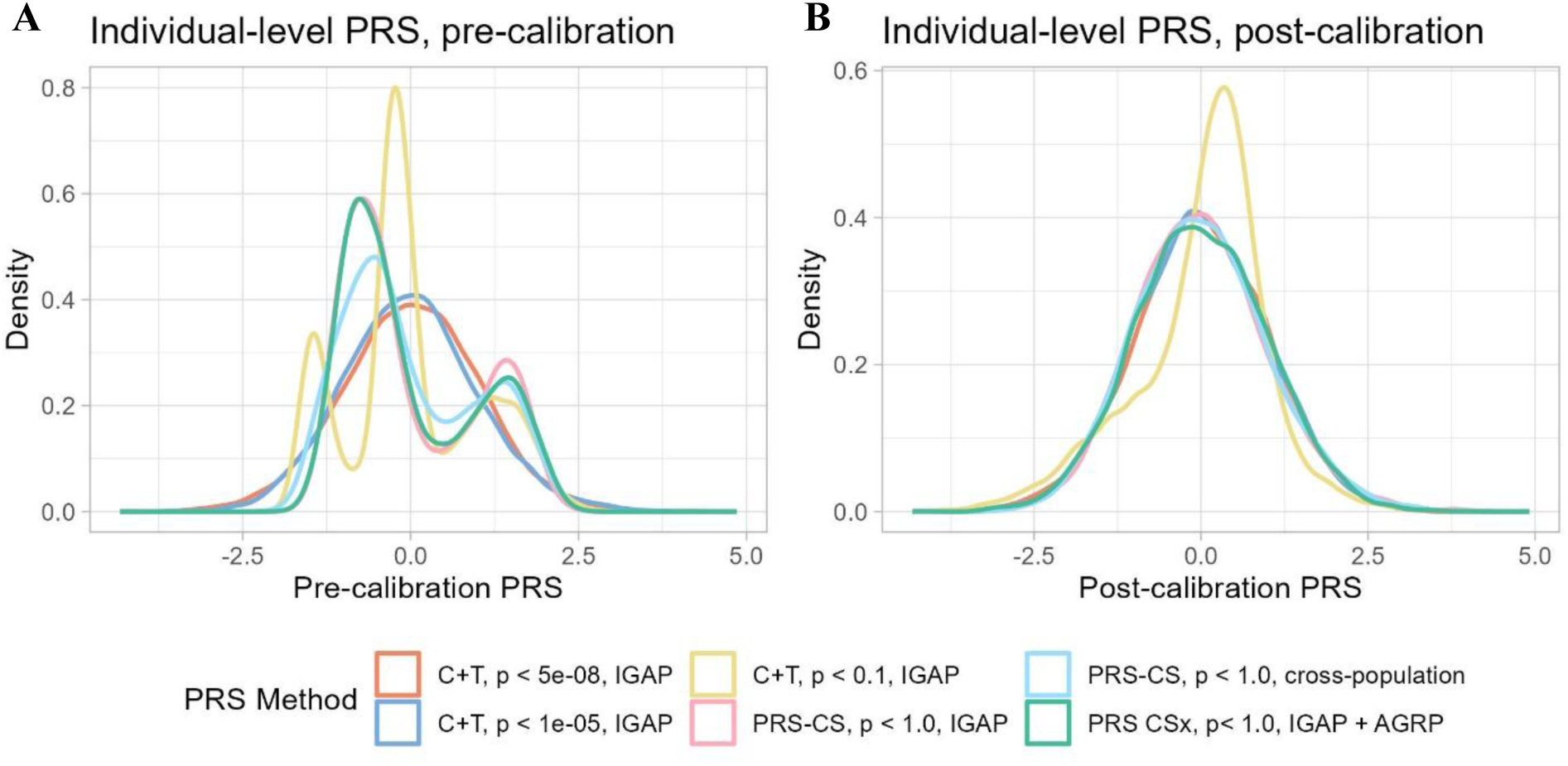
PRS distributions before and after calibration by principal components. Density plots show the distribution of risk scores for each PRS method across all participants. A) Distributions of scores that have been mean-standardized but not adjusted for principal components. B) Distributions of scores after standardization and principal component calibration.

### 3.3 Association between PRS and Incident Dementia

Univariate Cox proportional hazards models were fit to test the association of each PRS model with incident dementia. In the full sample, PRS derived from all models except for the C+T model with *p*-value threshold *<*0.1 were associated with incident dementia (**Figure 2**). The hazard ratio for the PRS constructed from the most stringent *p*-value threshold (*p <* 5e-08 C+T) (HR_5e-08_ = 1.21, 95% CI: 1.11-1.31, was higher than the other models, but this difference was not statistically significant (HR_1e-05_ = 1.12, 95% CI: 1.02-1.22; HR_0.01_ = 0.95, 95% CI: 0.87-1.04; HR_CS-EUR_ = 1.13, 95% CI: 1.05-1.22; HR_CS-CP_ = 1.14, 95% CI: 1.06-1.24; HR_CSx_ = 1.13, 95% CI: 1.04-1.23; **Figure 2**).

**Figure 2.**
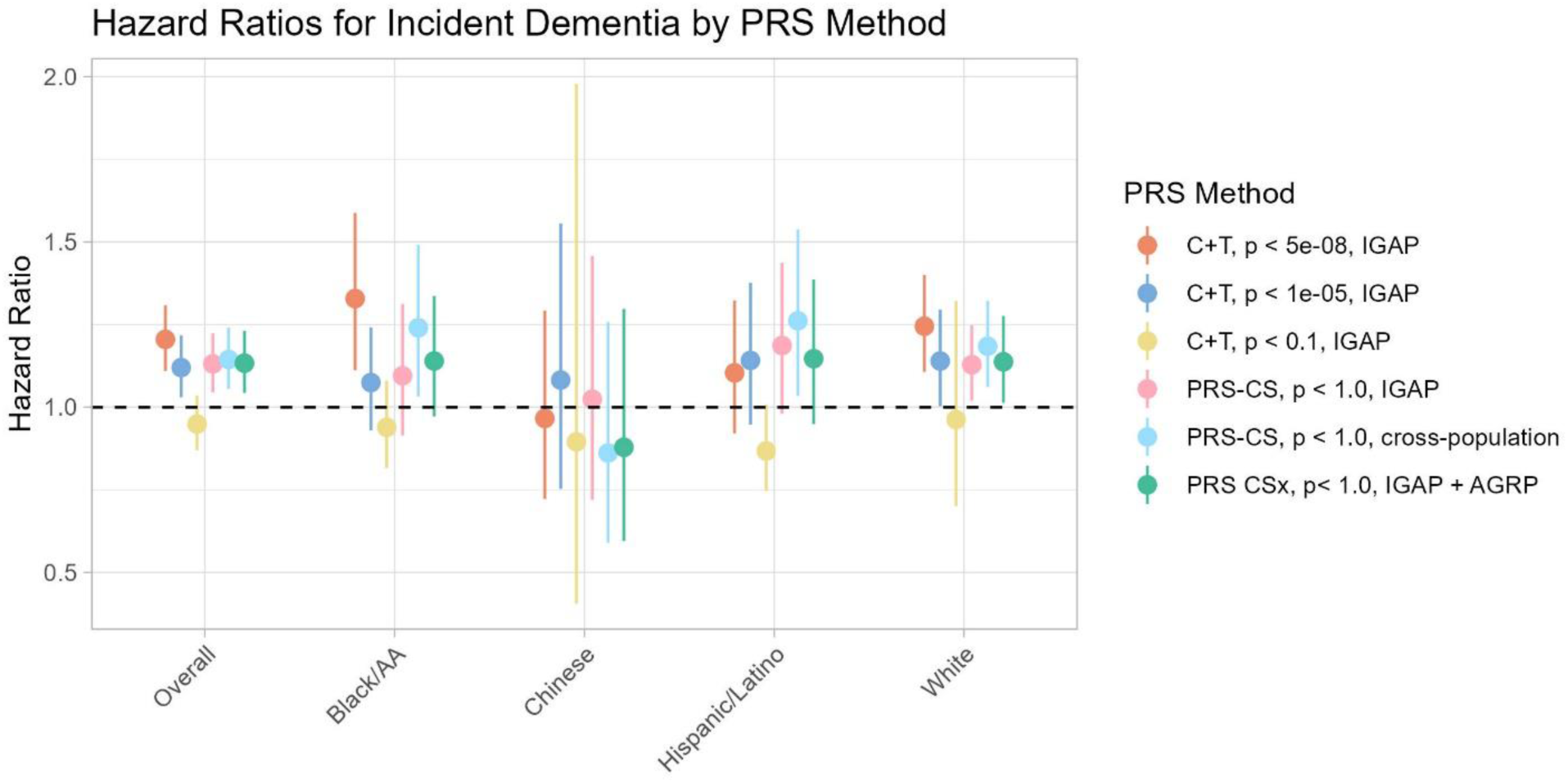
Association between PRS and incident dementia. The forest plots display the hazard ratio and confidence intervals for univariate models with the PRS as exposure and incident dementia outcome. Results are presented for all participants (Overall) and for groups stratified by self-reported race/ethnicity.

Among the race/ethnicity-stratified models, the cross-population PRS-CS model was associated with dementia in Black/African American (HR_AA_CS-CP_ = 1.24, 95% CI: 1.03-1.49), Hispanic/Latino (HR_HIS_CS-CP_ = 1.26, 95% CI: 1.03-1.54), and White groups (HR_WHI_CS-CP_ = 1.18, 95% CI: 1.06-1.32). The *p <* 5e-08 C+T model was associated with incident dementia among the African American/Black (HR_AA_5e-08_ = 1.33, 95% CI: 1.11-1.59) and White participants (HR_WHI_5e-08_ = 1.24, 95% CI: 1.11-1.40). No PRS were significantly associated with dementia in the Chinese participants, likely due to the limited number of observations (40 cases, 734 censored).

Under the assumption that model performance may be more dependent on genetic similarity to the GWAS sample than self-reported race/ethnicity, we also stratified MESA participants into tertiles of NFE-like ancestry. Among the low, intermediate, and high NFE-like groups, we found that in the high NFE-like group (n = 2,651), all PRS aside from the *p <* 0.1 C+T model were significantly associated with the hazard of dementia **(Supplementary Figure 3**). In contrast, no PRS was associated with the hazard of dementia in the intermediate NFE-like group (n = 1,256) regardless of approach, and only the most stringent C+T PRS*<* was associated with dementia in the low NFE-like group (n = 2,431, HR_lowNFE_5e-08_ = 1.26, 95% CI: 1.08-1.47).

As a sensitivity analysis, we fit univariate logistic regression models to assess the association between the PRS models and dementia case-control status. The results from the logistic regression model mirror findings from the Cox proportional hazards models. Among all MESA participants, the *p <* 5e-08 C+T model had stronger estimated association with dementia status compared to other models, but the difference was not statistically significant (OR_5e-08_ = 1.21, 95% CI: 1.11-1.32; OR _1e-05_ = 1.12, 95% CI: 1.02-1.22; OR _0.01_ = 0.95, 95% CI: 0.87-1.04; OR _CS-EUR_ = 1.15, 95% CI: 1.06-1.26; OR _CS-CP_ = 1.17, 95% CI: 1.07-1.27; OR _CSx_ = 1.16, 95% CI: 1.06-1.26; **Supplementary Figure 4**).

### 3.4 Assessing Model Performance

Model performance was evaluated using Harrell’s concordance index (C-index), with the highest value observed for the most stringent*<* C+T model (C_5e-08_ = 0.55, standard deviation (SD) = 0.01). Comparisons across models revealed that the inclusion of more SNPs in either C+T models using Bayesian approaches does not improve predictive accuracy (**Figure 3**). These findings were also supported by comparisons of model AUC (AUC_5e-08_ = 0.56, SD = 0.01, **Supplementary Figure 5**).

**Figure 3.**
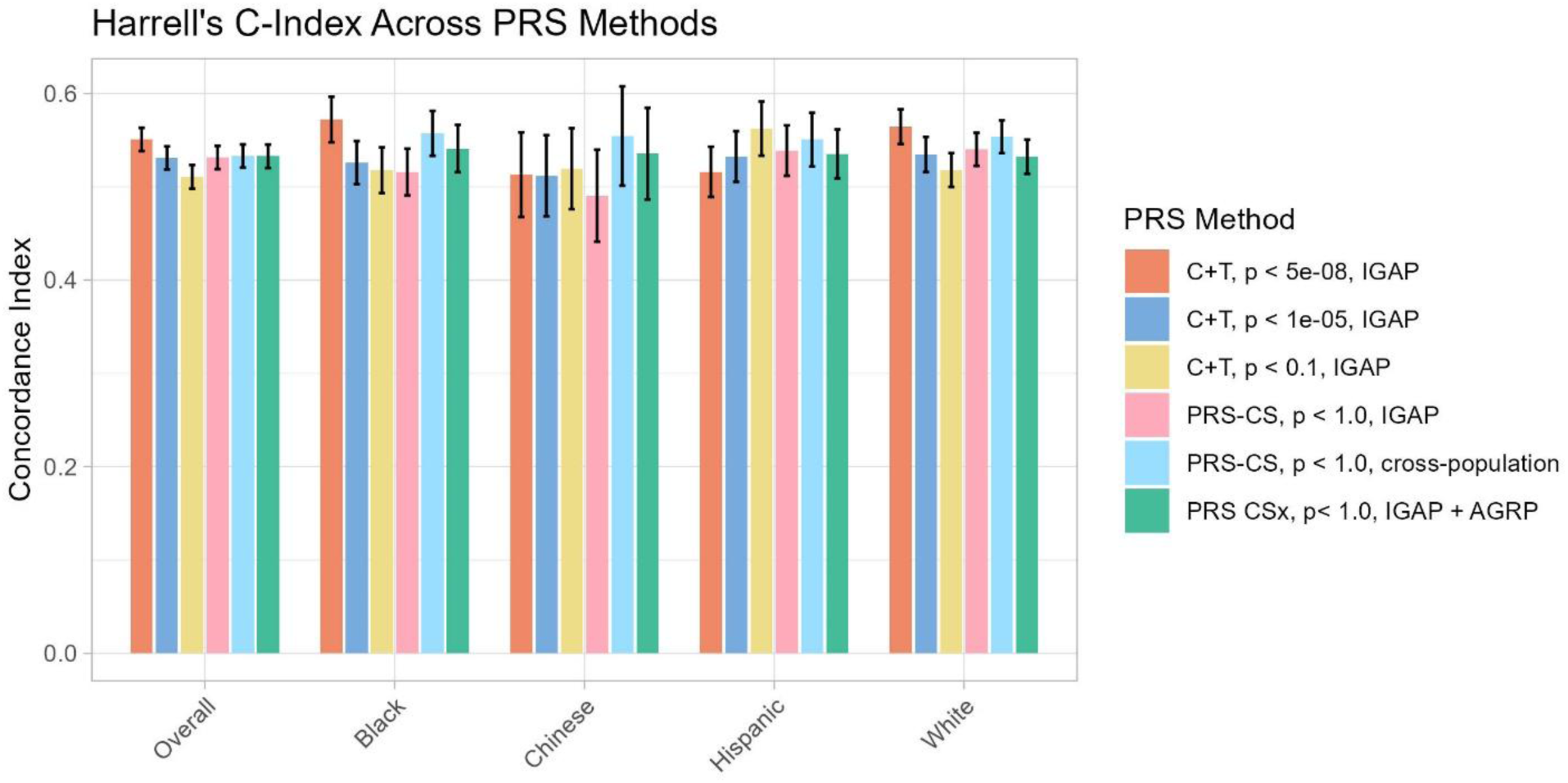
PRS predictive performance measured by Harrell’s Concordance Index. The bar plots show a comparison of prediction accuracy as measured by the concordance index or Harrell’s C. Error bars represent 95% confidence intervals.

In groups with low and high proportions of NFE-like ancestry, the *p <* 5e-08 C+T model had the highest C-index (C_lowNFE_5e-08_ = 0.57, SD = 0.02; C _highNFE_5e-08_ = 0.56, SD = 0.02, **Supplementary Figure 6**). In the group with intermediate NFE-like proportion, the PRS-CSx model had the best performance (C_midNFE_csx_ = 0.54, SD = 0.03) while the *p <* 5e-08 C+T model had the worst performance (C_midNFE_5e-08_ = 0.51, SD = 0.02).

### 3.5 Value added from prediction using PRS

Compared to a baseline model that included age, sex, and *APOE* genotype, the addition of PRS derived from *p <* 5e-08 and *p <* 1e-05 C+T models and the PRS-CSx model led to a marginal increase in C-index. The baseline model had a C-index of 0.84. In the *p <* 5e-08 C+T model, *p <* 1e05 C+T model, and PRS-CSx model, inclusion of the PRS significantly improved model fit (*p*_LRT_5e-08_ = 5e-05, *p*_LRT_1e05_ = 0.008, *p*_LRT_CSx_ = 0.001, **Table 3**). The remaining PRS models did not improve model fit.

**Table 3.** Value Added of PRS. We computed the added predictive value of each PRS method compared to baseline models that included sex, age, and *APOEɛ4* carrier status and used likelihood ratio tests to test the significance of improved model fit when including the PRS. This table shows the change in concordance (C-statistic) from adding PRS to the model and the *p*-value corresponding to the likelihood ratio test.

| PRS Model | Δ Concordance | *p*-value<sub>LRT</sub> |
| --- | --- | --- |
| C+T, *p* <5e-08 | 0.0015 | 5e-05* |
| C+T, *p* <1e-05 | 0.0007 | 0.008* |
| C+T, *p* <0.01 | 0.0001 | 0.903 |
| PRS-CS, IGAP | 0.0001 | 0.220 |
| PRS-CS, Cross-Pop | 0.0002 | 0.128 |
| PRS-CSx | 0.0014 | 0.001* |

## 4. Discussion

Improved prediction of Alzheimer’s disease and dementia is urgently needed for advancing research into novel treatment and prevention strategies. PRS are increasingly being used to assess genetic susceptibility for a wide spectrum of diseases, allowing for earlier identification of individuals at higher risk, but few studies have assessed the association of Alzheimer’s disease PRS for predicting dementia in diverse populations. Our study demonstrates that PRS models, even when excluding the *APOE* region, remain significantly associated with incident dementia in a multi-ancestry sample. We also found that including more SNPs does not improve predictive performance. A smaller panel of variants is more robustly associated with incident dementia across diverse groups than the larger models. However, even the best performing PRS models in our study have low predictive power (C-index *<* 0.6) and add only slight improvements to predictive models that include age, sex, and *APOE*. Early prediction of dementia will likely require integrating demographic and environmental information in addition to genetics.

Our comparisons also demonstrate the benefit of diversifying genomic studies of AD, adding to the growing calls for diversity across genetic research. The PRS-CS model using summary statistics from cross-population GWAS of AD consistently performed better than the PRS-CS model using summary statistics from the European-ancestry GWAS, despite the smaller sample size of the cross-population GWAS. This finding is consistent with previous work examining other non-dementia traits that demonstrated the superior performance of PRS derived from multi-ancestry GWAS meta-analyses compared to single-ancestry GWAS(Gunn et al., 2025). Of note, neither of the PRS-CS models outperformed the restrictive C+T model with 15 SNPs derived from the IGAP GWAS representing European ancestry. It’s likely that the restricted set of SNPs are more likely to tag regions that have true biological impact on risk of disease development.

PRS are least predictive in individuals with high amounts of genetic admixture – those in the intermediate NFE-like proportion group. We observed that PRS-CSx performed best in the group with intermediate proportions of NFE-like ancestry. This aligns with previous findings that have observed increased predictive performance in admixed groups when using a linear combination of summary statistics that combine ancestry-specific effect sizes(Bitarello & Mathieson, 2020). Nevertheless, the PRS-CSx performance in this group remained lower than the best performing models in the high NFE-like group and the PRS derived from PRS-CSx was not associated with incident dementia in this group (**Supplementary Figure 3**). The limited association could be due to the relatively small sample size of the African Genome Resources Panel GWAS and lack of GWAS information from studies with substantial AMR ancestry.

Our study has several limitations. Incident dementia cases were ascertained as “possible dementia” from hospitalization and death records using ICD codes, and this method likely underestimates the true incidence because a portion of individuals living with dementia will not be hospitalized or have an alternative listed cause of death. ICD based possible dementia is also not specific for AD. However, the external validation of electronic health records by physician review and the association of both *APOE* genotype and our PRS with incident dementia suggests that dementia cases are true positives(Fujiyoshi et al., 2017). Furthermore, there are larger GWAS of dementia-by-proxy phenotypes conducted in European ancestry samples that provide a larger pool of SNPs considered to be significantly associated with parental history of dementia. Due to the sub-optimal dementia adjudication of our target data, we chose to prioritize depth of phenotyping over sample size. We also limited our summary statistics to variants identified in GWAS and, therefore, did not consider rare variants that may have large effects on AD. In addition, novel polygenic risk approaches are constantly being developed and we are unable to test all possible methods. Instead, we selected methods that are shown to perform well in the absence of individual level validation data due to the lack of datasets that match the diversity of our target sample and are not included in the GWAS from which the summary statistics are derived. Finally, while Chinese participants were included in our study, the sample size was too small to detect differences in performance across PRS models and none of the models resulted in PRS associated with dementia in this subgroup.

While our findings show that current PRS models have modest predictive value, future AD GWAS in diverse populations have the potential to enhance their predictive power.

Furthermore, the utility of PRS may extend beyond estimating the overall likelihood of disease development. Future research should focus on how PRS can be used to identify differences in disease pathogenesis and clinical trajectories using adjudicated cases of dementia and specifically AD in diverse populations. Incorporation of rare variants and leveraging functional annotation to develop pathway specific scores will further enhance the precision and translational value of PRS. For now, it seems there is no tradeoff between simplicity and accuracy; a simple C+T approach with only the most highly significant SNPs offers comparable, if not superior, prediction of dementia in populations with diverse ancestry.

## Data Availability

Data produced in the present study are available upon reasonable request to the authors

## Acknowledgements

This research was supported by NIA F99AG079792 and NIA K01 AG071689. MESA and the MESA SHARe projects are conducted and supported by the National Heart, Lung, and Blood Institute (NHLBI) in collaboration with MESA investigators. Support for MESA is provided by contracts 75N92020D00001, HHSN268201500003I, N01-HC-95159, 75N92020D00005, N01-HC-95160, 75N92020D00002, N01-HC-95161, 75N92020D00003, N01-HC-95162, 75N92020D00006, N01-HC-95163, 75N92020D00004, N01-HC-95164, 75N92020D00007, N01-HC-95165, N01-HC-95166, N01-HC-95167, N01-HC-95168, N01-HC-95169, UL1-TR-000040, UL1-TR-001079, and UL1-TR-001420, UL1TR001881, DK063491, R01HL105756, and R01AG058969. Funding for SHARe genotyping was provided by NHLBI Contract N02-HL-64278. Genotyping was performed at Affymetrix (Santa Clara, California, USA) and the Broad Institute of Harvard and MIT (Boston, Massachusetts, USA) using the Affymetrix Genome-Wide Human SNP Array 6.0. The authors thank the other investigators, the staff, and the participants of the MESA study for their valuable contributions. A full list of participating MESA investigators and institutes can be found at http://www.mesa-nhlbi.org.

---

**Supplementary Figure 1.**
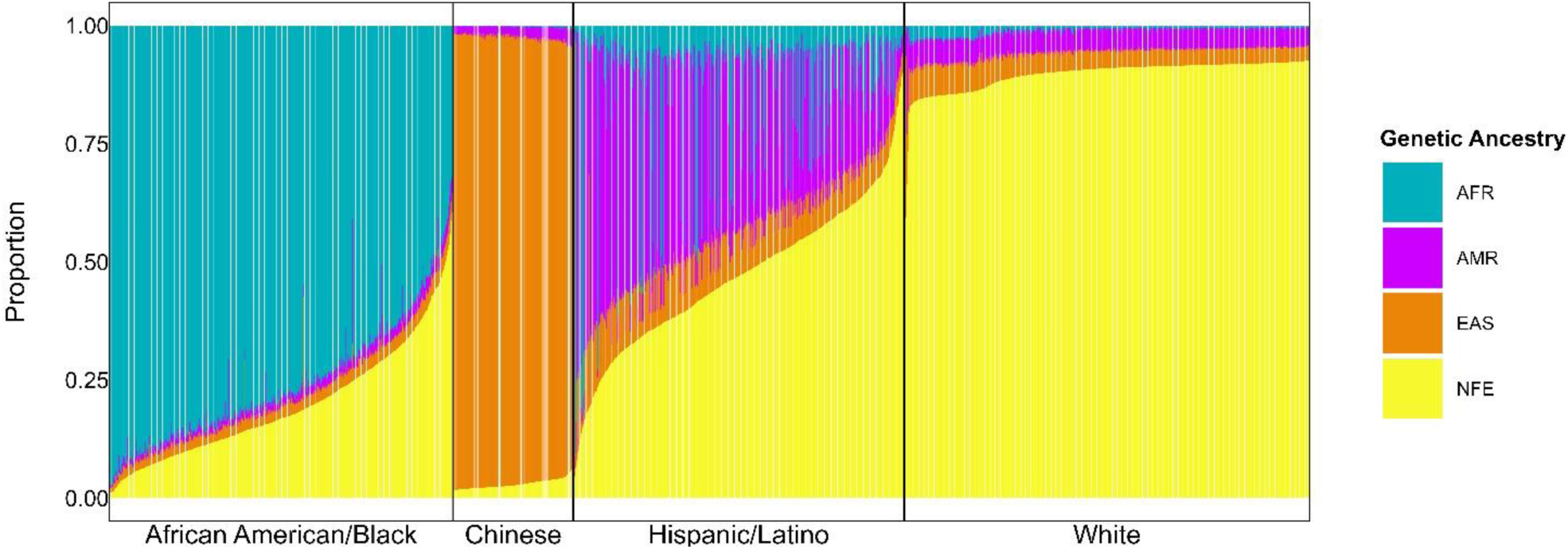
Global Ancestry Proportions for MESA participants. This plot represents the proportion of African (AFR), Amerindian (AMR), East Asian (EAS) and Non-Finnish European (NFE) for each participant, separated by self-reported race/ethnicity. Each participant represents one column. Reference samples are from gnomAD v3.1.

**Supplementary Figure 2.**
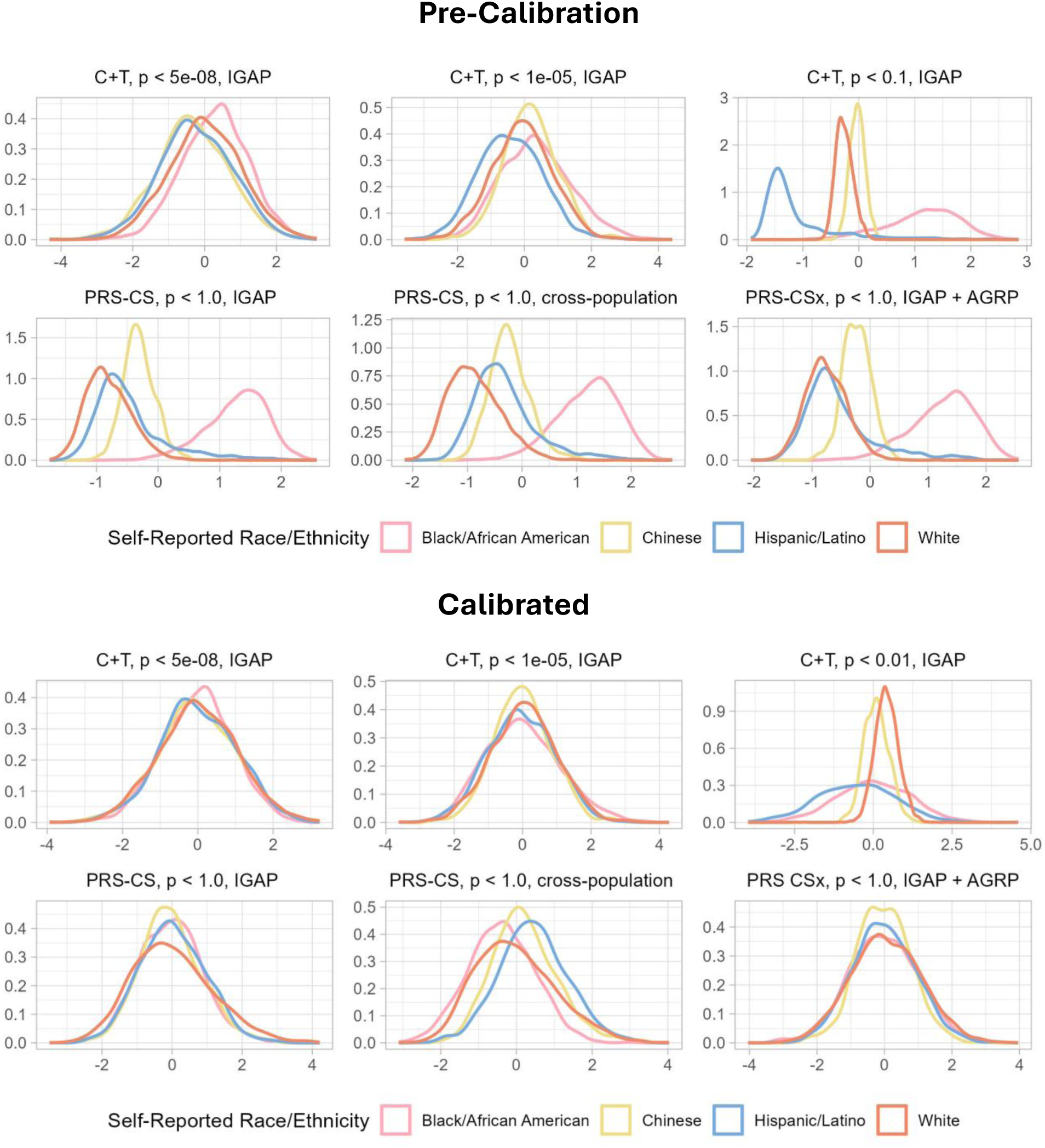
Polygenic risk score distributions by self-reported race/ethnicity, pre- and post-calibration by principal components.

**Supplementary Figure 3.**
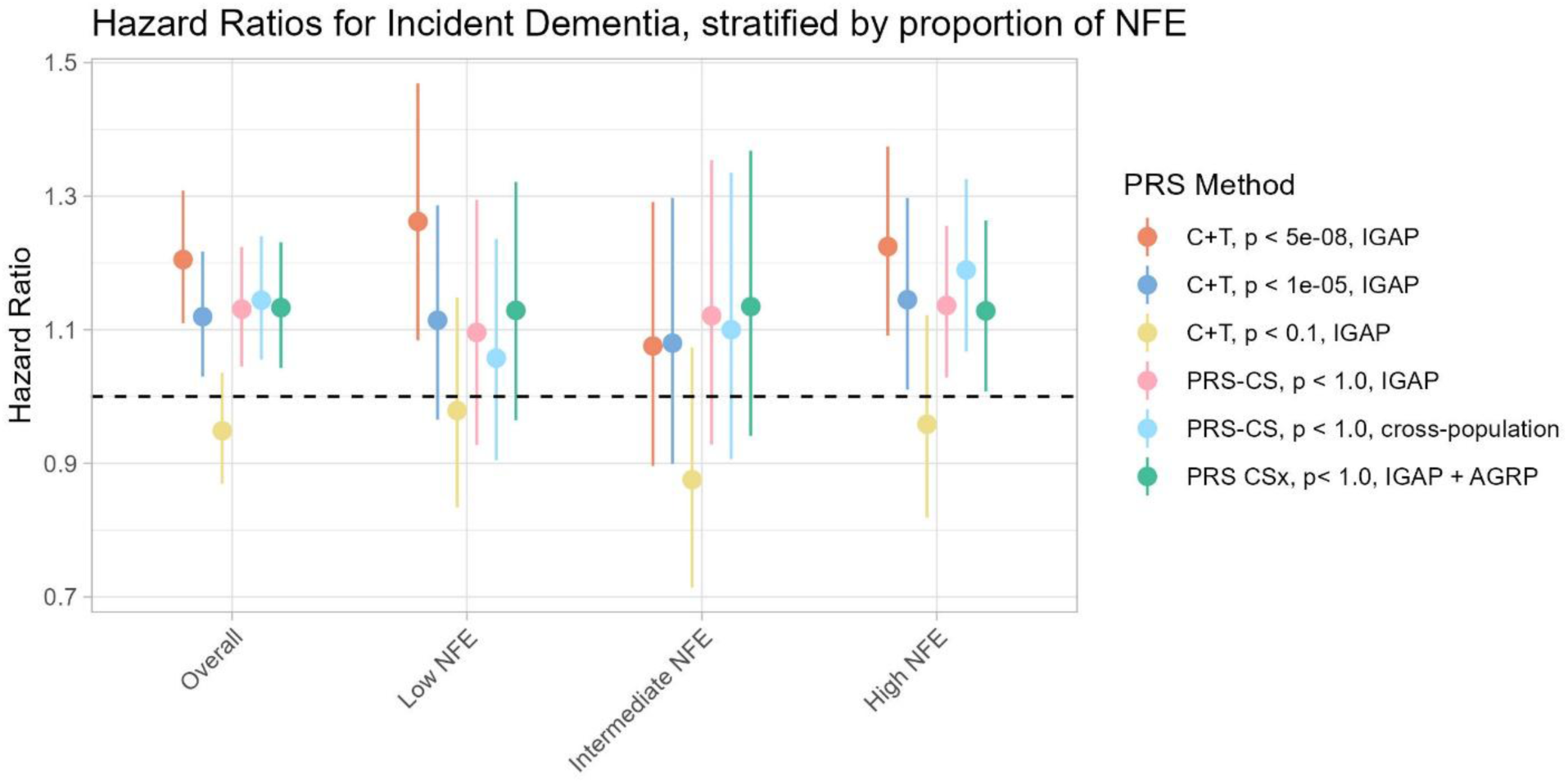
Association between adjusted PRS and incident dementia stratified by proportion of NFE ancestry. *NFE = proportion of ancestry similar to 1000 Genomes non-Finnish European references.

**Supplementary Figure 4.**
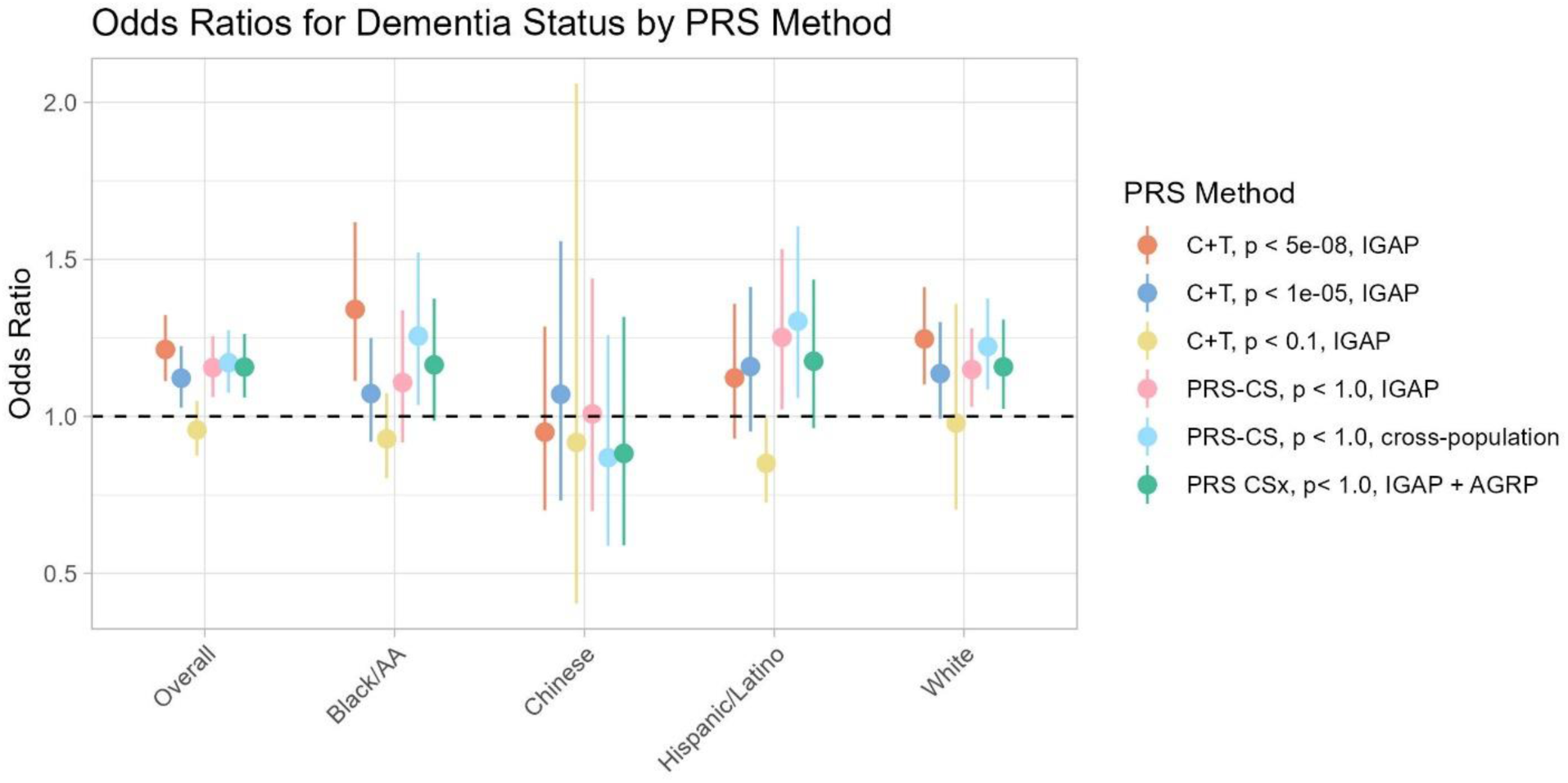
Association between adjusted PRS and dementia case-control status.

**Supplementary Figure 5.**
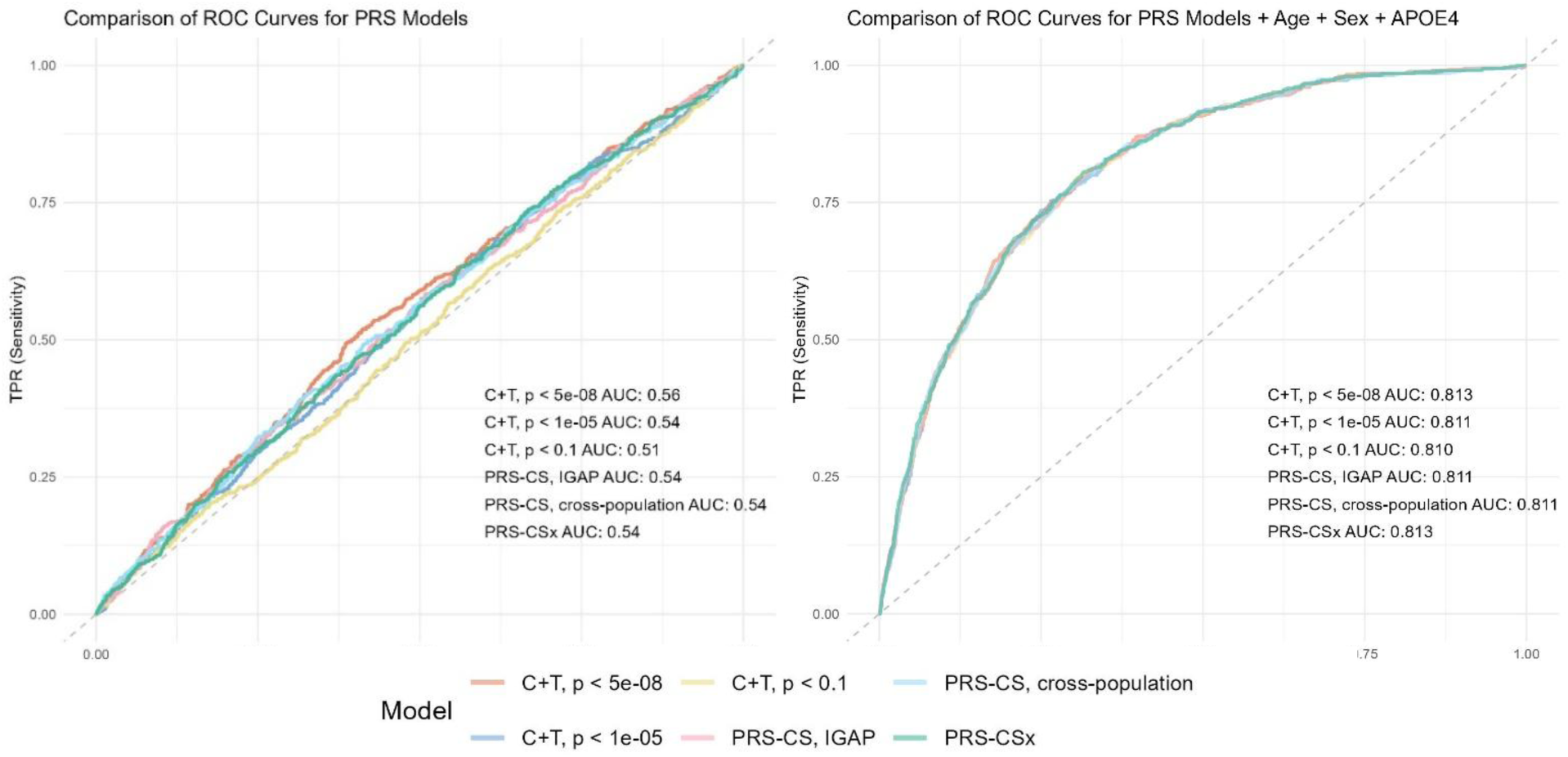
PRS predictive performance measured by AUC. The left-hand panel represents the predictive performance of a univariate model where the PRS alone are used to predict dementia status. The right-hand panel represents the predictive performance of a model that includes the PRS alongside age, sex, and *APOEe4* dosage.

**Supplementary Figure 6.**
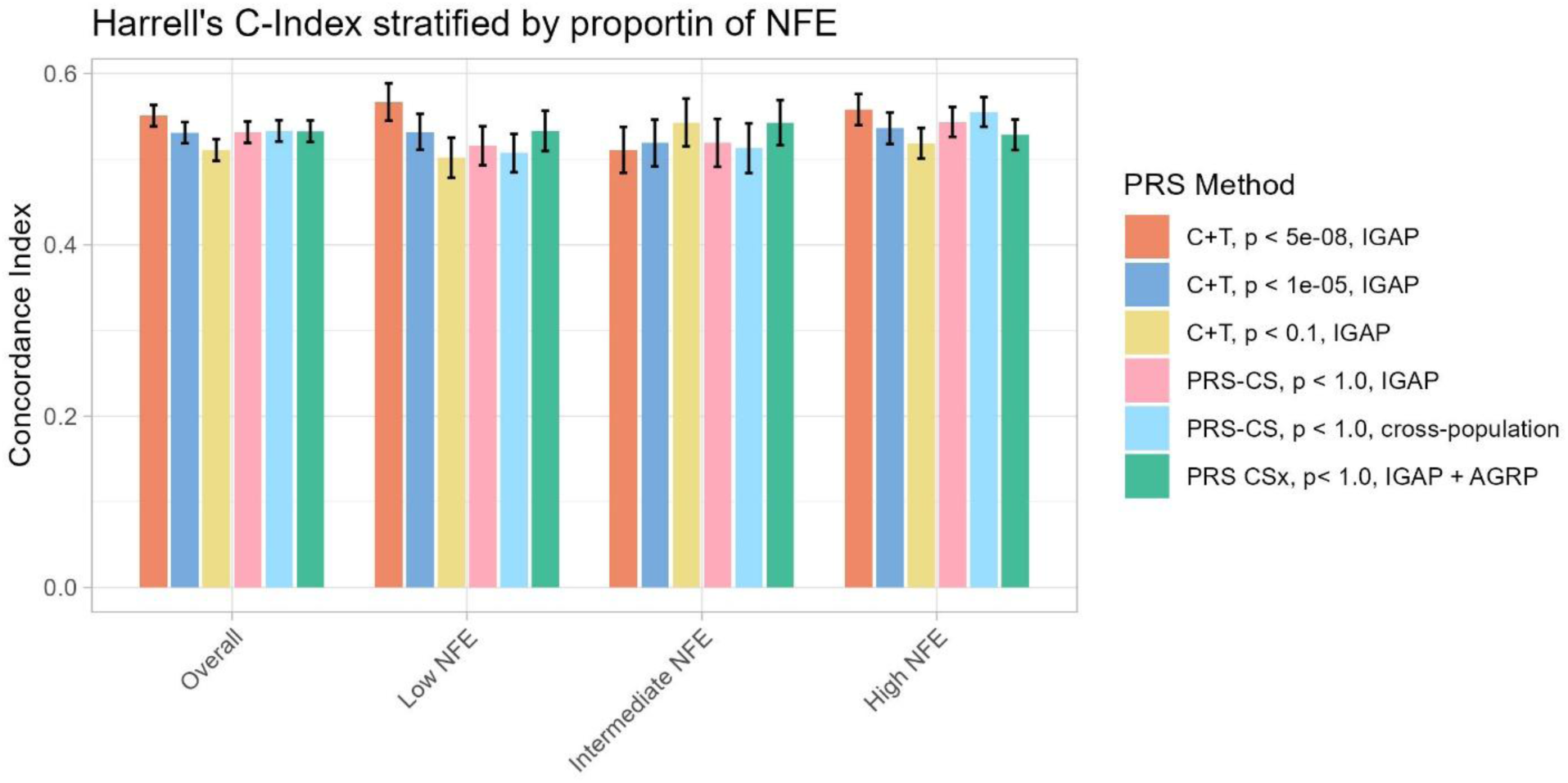
PRS predictive performance comparisons stratified by proportion of NFE-like ancestry. Comparisons of PRS predictive performance as measured by Harrell’s C across tertiles of NFE-like ancestry. *NFE = proportion of ancestry similar to 1000 Genomes non-Finnish European references.

